# The lower-airway microbiome and metabolome in preterm infants: Identifying potential predictive biomarkers of bronchopulmonary dysplasia

**DOI:** 10.1101/2020.09.02.20186601

**Authors:** Qi Xu, Jialin Yu, Junli He, Qi Tan, Yu He

## Abstract

The lower-airway microbiome may influence the pathogenesis of lung disease. Bronchopulmonary dysplasia (BPD) is a serious morbidity associated with preterm birth that may be influenced by lower-airway microbial or metabolic alterations. This study used16S rRNA gene sequencing, metabolomic analyses, and the Kyoto Encyclopedia of Genes and Genomes (KEGG) database to investigate the lower-airway microbiome and metabolome in a cohort of preterm infants with mild, moderate, or severe BPD or no BPD. Differences in the diversity and composition of the infants’ lower airway microbiota, as well as metabolic status, were initially observed, but became less pronounced at 7 days of life. Decreased diversity of the lower-airway microbiome, increased abundance of *Stenotrophomonas*, and increased level of sn-glycerol 3-phosphoethanolamine were associated with increased BPD severity, and have potential as predictive biomarkers for BPD. *Stenotrophomonas* may contribute to the development of BPD and influence the composition of the lower-airway microbiome through its metabolite, sn-glycerol 3-phosphoethanolamine. These findings provide novel insights into the lower-airway microbiome and its role in BPD.

## 1. Introduction

Bronchopulmonary dysplasia (BPD) is a serious morbidity associated with preterm birth that affects an estimated 50% of infants born at < 28 weeks of gestation [1]. Infants with BPD have an increased risk of mortality during the first year. Those who survive may suffer long-term pulmonary impairment and abnormal neurodevelopment, which can result in substantial healthcare resource utilization and cost [2]. Risk factors for BPD include, but are not limited to, gestational age at birth, impaired growth for gestational age, low infant birth weight, infectious exposures, barotrauma, oxygen exposure, and environmental cigarette smoke [3].

Evidence from epidemiological data, clinical data, and animal models indicate a key role for the microbiome in lung disease [4–6], and indicate that the lower-airway microbiome is altered in multiple respiratory disorders [7, 8]. Some reports show that the lower-airway microbiome is present at birth, and microbial dysbiosis may be associated with BPD [9, 10]. However, by now the related research is rare and the present results differ from each other, so it needs further research. What’s more, the mechanisms by which the microbiome alterations lead to BPD have never been addressed. Gut microbiota is associated with a variety of human diseases through metabolites [11]. We hypothesized that lower-airway microbial metabolism plays a role in the pathogenesis of BPD.

In this prospective observational cohort study, tracheal aspirates (TA) were collected during mechanical ventilation of infants to investigate 1) the lower-airway microbiome at birth (Day 1) and on Day 7 after birth; 2) the lower-airway metabolomic signatures at birth and on Day 7 after birth; and 3) the relationship between differential metabolites and specific bacteria among infants with severe BPD, moderate BPD, mild BPD, and no BPD.

## 2. Methods

This prospective observational cohort study was conducted at the Neonatal Intensive Care Unit of the Children’s Hospital of Chongqing Medical University between October 2017 and July 2018. The Institutional Review Board of Chongqing Medical University approved the protocol. Informed consent was received from the parents or guardians of all participants. The study was performed in accordance with approved guidelines.

### 2.1 Patient population and clinical data collection

Infants born at < 34 weeks gestation that underwent endotracheal intubation and mechanical ventilation in the first 24 h of life and remained intubated until at least Day 7 after birth were included in this study. Exclusion criteria were: 1) clinical evidence of congenital heart disease (except patent ductus arteriosus [PDA], patent foramen ovale [PFO] or atrial septal defect [ASD] *<* 1cm, or ventricular septal defect [VSD] *<* 2mm if known prior to enrollment); 2) lethal congenital abnormality; 3) congenital sepsis; 4) evidence of pulmonary hypoplasia; or 5) futile cases (anticipated death prior to hospital discharge) [10].

Infants were divided into four groups stratified by the diagnosis and severity of BPD: severe BPD, moderate BPD, mild BPD, and no BPD. BPD was diagnosed based on the need for supplemental oxygen at 28 days of age [12, 13]. BPD status and severity was assessed at 36 weeks postmenstrual age according to the National Institutes of Health workshop definition [13]. Late onset sepsis was defined as a positive blood culture after 72 h of life.

Clinical data were collected from a review of electronic medical records at study enrollment and during hospitalization. Information on maternal history, delivery, and clinical assessments was recorded.

### 2.2 Sample collection

Tracheal aspirates (TA) were collected during mechanical ventilation at birth (Day 1) and on Day 7 after birth according to a previously published protocol [13–15]. Briefly, 0.5 ml of sterile isotonic saline was instilled into the infants’ endotracheal tubes. Infants were manually ventilated through their endotracheal tube for three breaths using a bag-mask, and fluid was suctioned into a sterile mucus trap [14]. Samples were divided into 2 aliquots for extraction of bacterial DNA or metabolomics research and frozen at –80°C until further processing.

### 2.3 Isolation of microbial DNA, creation of 16S V4 amplicon library, and DNA sequencing

Microbial genomic DNA from each sample was isolated and purified. The V4 region of the 16S rRNA gene from the microbial DNA was amplified using a polymerase chain reaction (PCR) with unique bar coded primers to create an “amplicon library” [16]. The library was sequenced using the Illumina MiSeq platform and subsequently quantified (KAPA Library Quantification Kit KK4824), according to the manufacturer’s instructions.

### 2.4 Profiling of 16S rRNA gene sequencing data

Raw sequences were processed with the Quantitative Insights into Microbial Ecology (QIIME) 1.8.0 pipeline1.The concat function was used to combine reads into tags according to an overlapping relationship. Reads from each sample were separated with barcodes, and low quality reads were removed. Processed tags were clustered at 97% similarity into operational taxonomic units (OTUs). Taxonomy was assigned to OTUs by matching to the Greengenes database (Release 13.8)2. Alpha diversity analyses (Shannon index) and beta diversity analyses (principal coordinate analysis [PCoA]) were performed.

### 2.4 Metabolomics analysis based on UHPLC-Q-TOF/MS

TA samples were analyzed using an ultra-high-performance liquid chromatography (UHPLC) system (1290 Infinity LC, Agilent Technologies) coupled to a quadrupole time-of-flight mass spectrometer (AB Sciex TripleTOF 6600) at Shanghai Applied Protein Technology Co., Ltd.

Samples were thawed at 4°C and 100 μL aliquots were mixed with 400 μL of cold methanol/acetonitrile (1:1, v/v) to remove the protein. After centrifuging for 15 min (14000g, 4 °C), the supernatant was dried in a vacuum centrifuge. For liquid chromatography-mass spectrometry (LC-MS), samples were dissolved in 100 μL acetonitrile/water (1:1, v/v). Pooled quality control (QC) samples were used to monitor the stability and repeatability of instrument analysis. The QC samples were inserted regularly and analyzed in every 5 samples.

Raw LS electrospray ionization (ESI) MS data were converted into m/z format and analyzed for non-linear retention time (RT) alignment, peak detection, and filtration.

### 2.5 Profiling of metabolomics data

Processed data were normalized to total peak intensity, imported into SIMCA-P (version 14.1, Umetrics, Umea, Sweden), and analyzed using Pareto-scaled principal component analysis (PCA) and orthogonal partial least-squares discriminant analysis (OPLS-DA). The variable importance in the projection (VIP) value for each variable in the OPLS-DA model was calculated to indicate its contribution to the classification. One-way ANOVA was used to determine the significance of each metabolite with a VIP value > 1. P< 0.05 were considered statistically significant. Discriminatory metabolites within the data set were visualized as heat maps, which were generated using a hierarchical clustering algorithm. Molecules associated with significant changes were searched against the Kyoto Encyclopedia of Genes and Genomes (KEGG) database (http://www.genome.jp/kegg/pathway.html).

### 2.6 Statistical analysis

Statistical analysis was performed using SPSS version 22.0 for Windows (SPSS Inc., USA). Normally distributed data are expressed as mean ± SD; non-normally distributed data are expressed as median and interquartile range (IQR). Between group differences were analyzed with Fisher’s Exact test for categorical variables and Kruskal-Wallis test for continuous variables after subsampling. Pairwise comparison was performed with White’s non-parametric t-test. Correlations between microbiome–related metabolites and bacterial species were evaluated using Pearson’s correlation coefficient. *P* < 0.05 was considered statistically significant.

## 3.0 Results

### 3.1 Demographic and clinical characteristics of the enrolled patients

This study included 30 premature infants divided into 4 groups, including 10 infants with severe BPD, 5 infants with moderate BPD, 10 infants with mild BPD, and 5 infants with no BPD. The demographic and clinical characteristics of the included infants are shown in **Table 1**. There were no significant differences in the demographic and clinical characteristics between the groups, except for number of hours of mechanical ventilation and number of days of antibiotics.

**Table 1.**
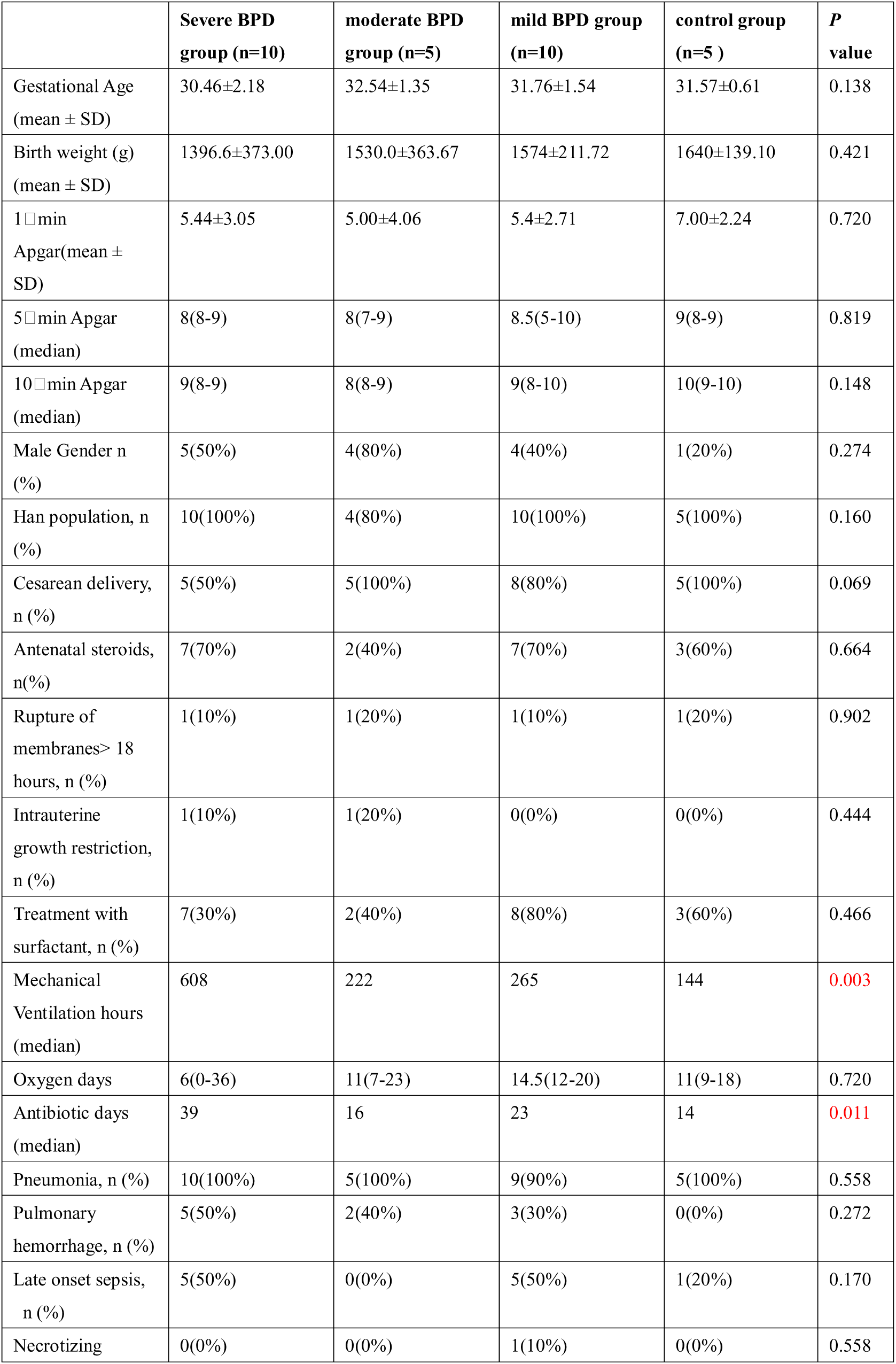
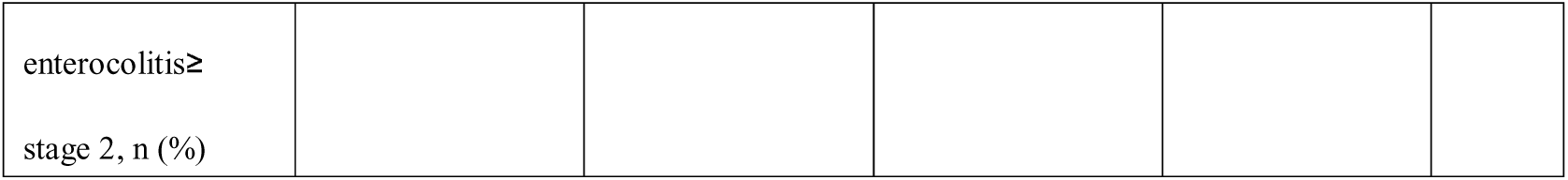
Demographic and clinical characteristics of the included patients.

### 3.2 Diversity and composition of the lower-airway microbiome

The Shannon index was significantly lower at birth (Day 1) (*P* = 0.0019; **Figure 1a**) and on Day 7 after birth (*P* = 0.016; **Figure 1b**) in infants with BPD compared to no BPD. The difference was more pronounced on Day 1 and was negatively correlated with the severity of BPD. Principal coordinates analysis (PCoA) showed a significant difference in the bacterial composition of the lower-airway microbiome at birth (Day 1) between the four groups (**Figure 1c**), and a less distinct difference on Day 7 after birth (**Figure 1d**).

**Figure 1.**
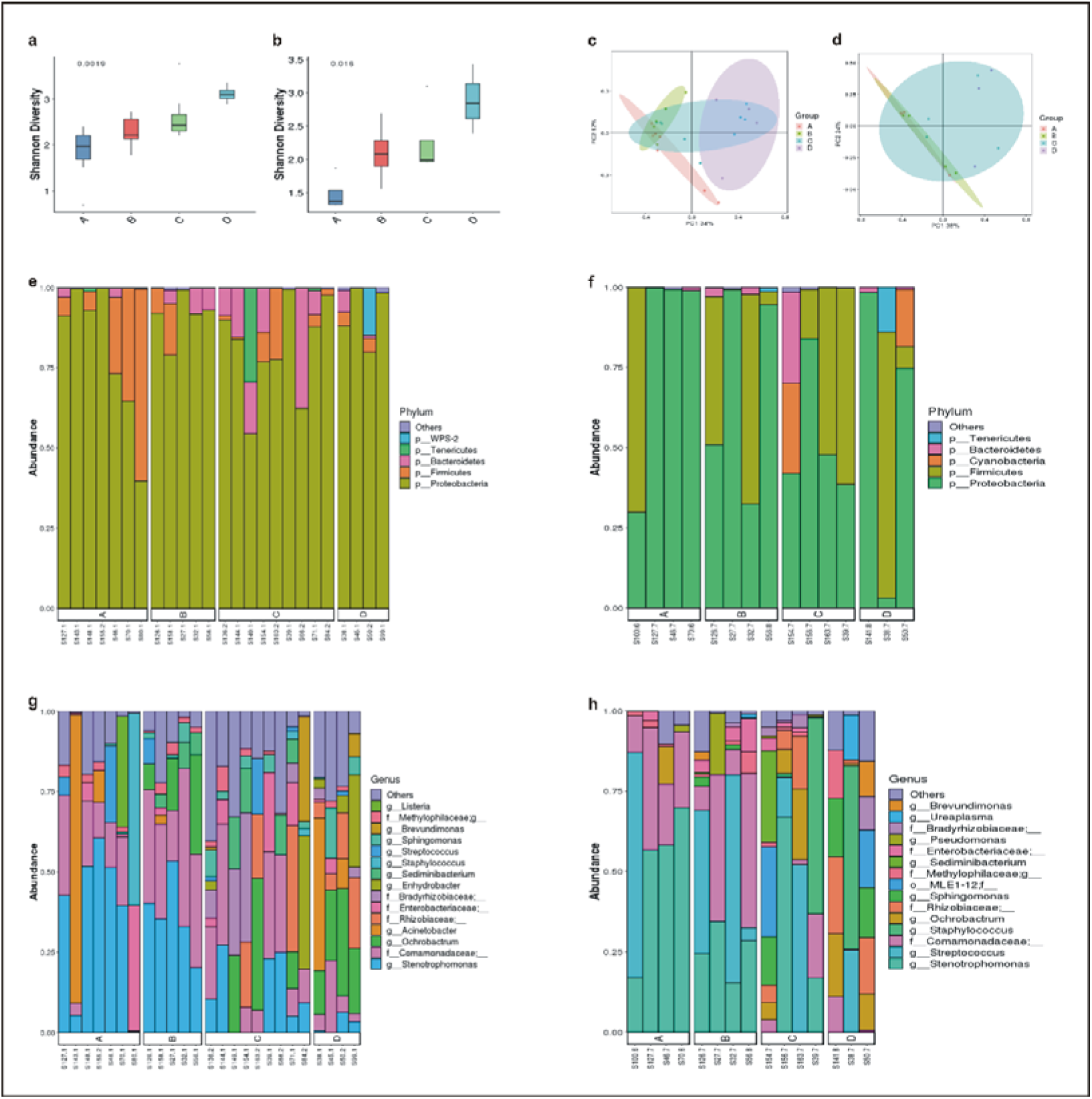
Diversity and composition of the lower-airway microbiome. (a) Shannon index at birth (Day 1); (b) Shannon index on Day 7 after birth (a greater Shannon index is indicative of higher microbial diversity); (c) Principal coordinate analysis (PCoA) of microbial communities at birth (Day 1) (d) PCoA of microbial communities on Day 7 after birth (samples located close to each other have similar microbial compositions, samples at distant locations have distinct microbial compositions); (e-f) Relative abundance of bacterial phyla (birth [Day 1], e; 7 days after birth, f). (g-h) Relative abundance of bacterial genera (birth [Day 1], g; 7 days after birth, h). (A, severe BPD [n = 10]; B, moderate BPD [n = 5]; C, mild BPD [n = 10]; D, no BPD [n = 5]).

At the phylum level, *Proteobacteria* was dominant in the lower-airway microbiome of all infants at birth (Day 1) (**Figure 1e**) and on Day 7 after birth (**Figure 1f**), and there were no significant differences in the composition of the lower-airway microbiome between the four groups. At the genus level, the composition of the lower-airway microbiome was significantly different between groups at birth (Day 1). *Stenotrophomonas* was more abundant in infants with BPD compared to no BPD, and abundance was positively correlated with the severity of disease (*P* < 0.05) (**Figure 2a**). Findings on Day 7 after birth were similar, but not statistically significant (*P* = 0.064) (**Figure 2b**).

**Figure 2.**
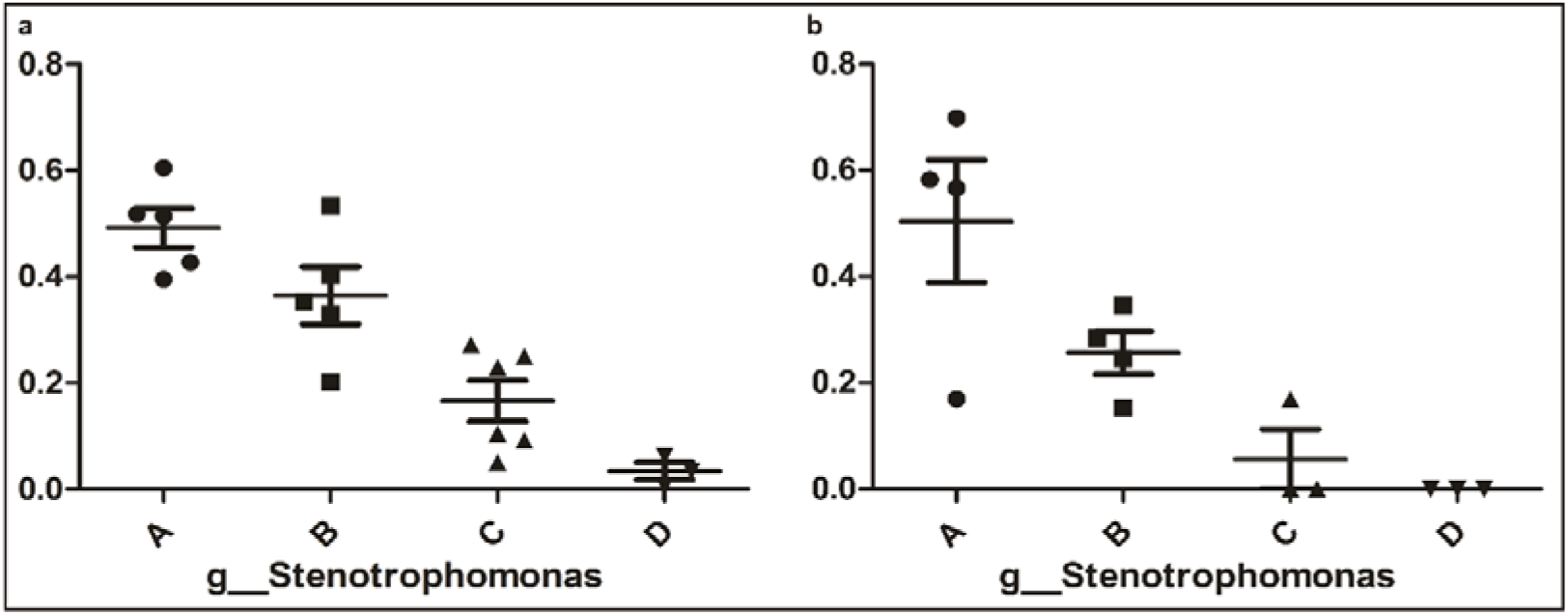
Abundance of *Stenotrophomonas*. (a) The abundance of *Stenotrophomonas* at birth (Day 1), *P* < 0.05; (b) the abundance of *Stenotrophomonas* on Day 7 after birth, *P* = 0.064. (A, severe BPD [n = 10]; B, moderate BPD [n = 5]; C, mild BPD [n = 10]; D, no BPD [n = 5]).

### 3.3 Metabolomic analysis of tracheal aspirates (TA)

TAs were subjected to LC/MS analysis in positive ion mode (ES+) and negative ion mode (ES-).

Principle component analysis (PCA) was performed to reduce dimensionality in the dataset (**Figure 3a, b**). Hierarchical clustering heat maps visualized patterns in molecular data across groups (**Figure 3c-f**). There were significant differences in 63 metabolites, including 23 in ES- and 40 in ES+ (Figure 3c,d), between the four groups at birth (Day 1), and 29 metabolites, including 11 in ES- and 18 in ES+, on Day 7 after birth (**Figure 3e-f**). Among these metabolites, sn-glycerol 3-phosphoethanolamine was positively correlated with BPD severity at birth (Day 1) (**Figure 4**), but not on Day 7 after birth.

**Figure 3:**
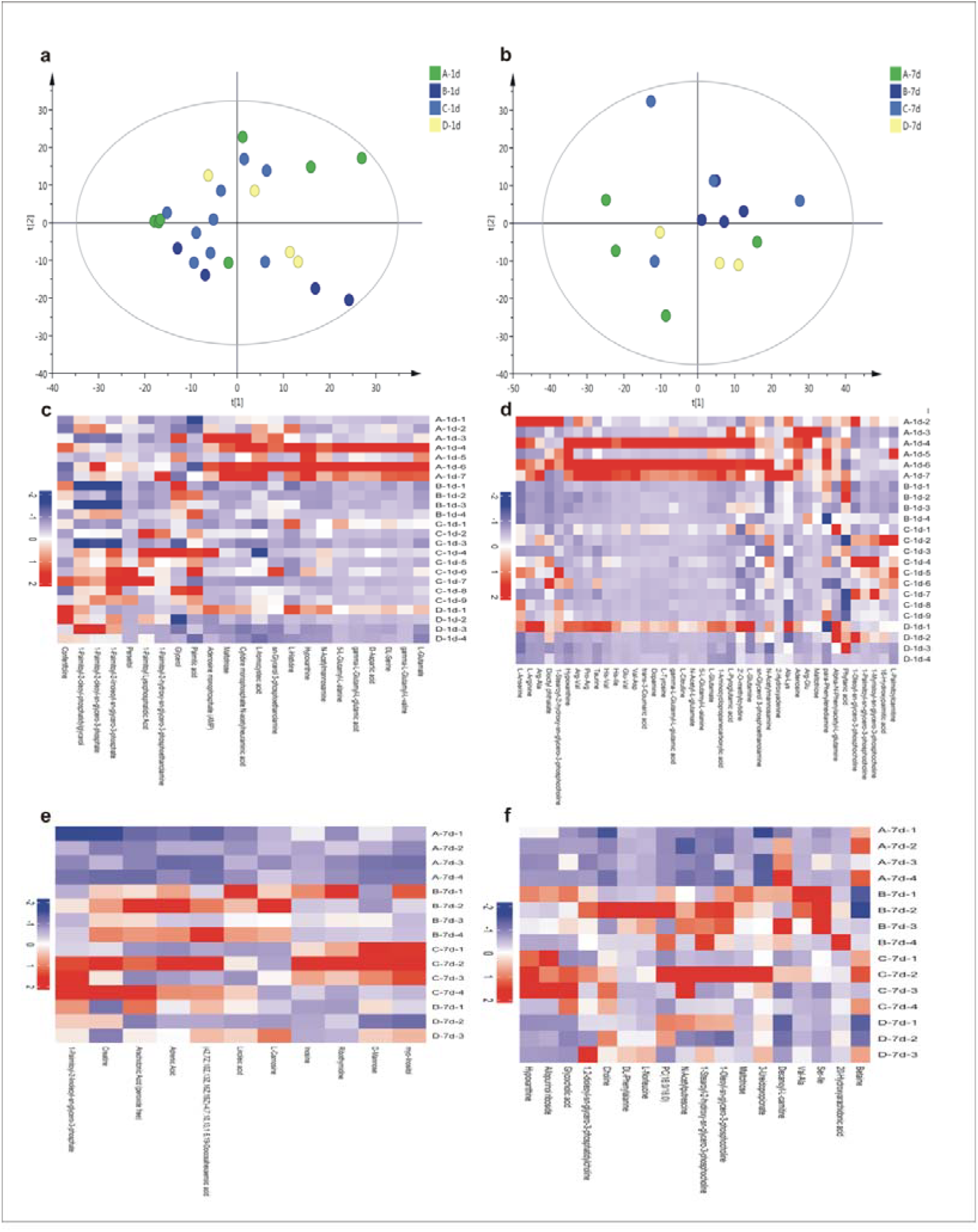
Metabolic profiles. (a-b) Principal Component Analysis (PCA) based on the metabolic profiles in sputum samples (birth [Day 1], a; 7 days after birth, b); (c-f) Hierarchical clustering heat maps showing patterns in molecular data across groups. The relative amounts of the 86 compounds were transformed into Z scores (birth [Day 1] ES-, c; ES+, d; 7 days after birth ES-, e, ES+, f). (A, severe BPD [n = 10]; B, moderate BPD [n = 5]; C, mild BPD [n = 10]; D, no BPD [n = 5]). ES+: positive ion mode, ES-: negative ion mode)

**Figure 4:**
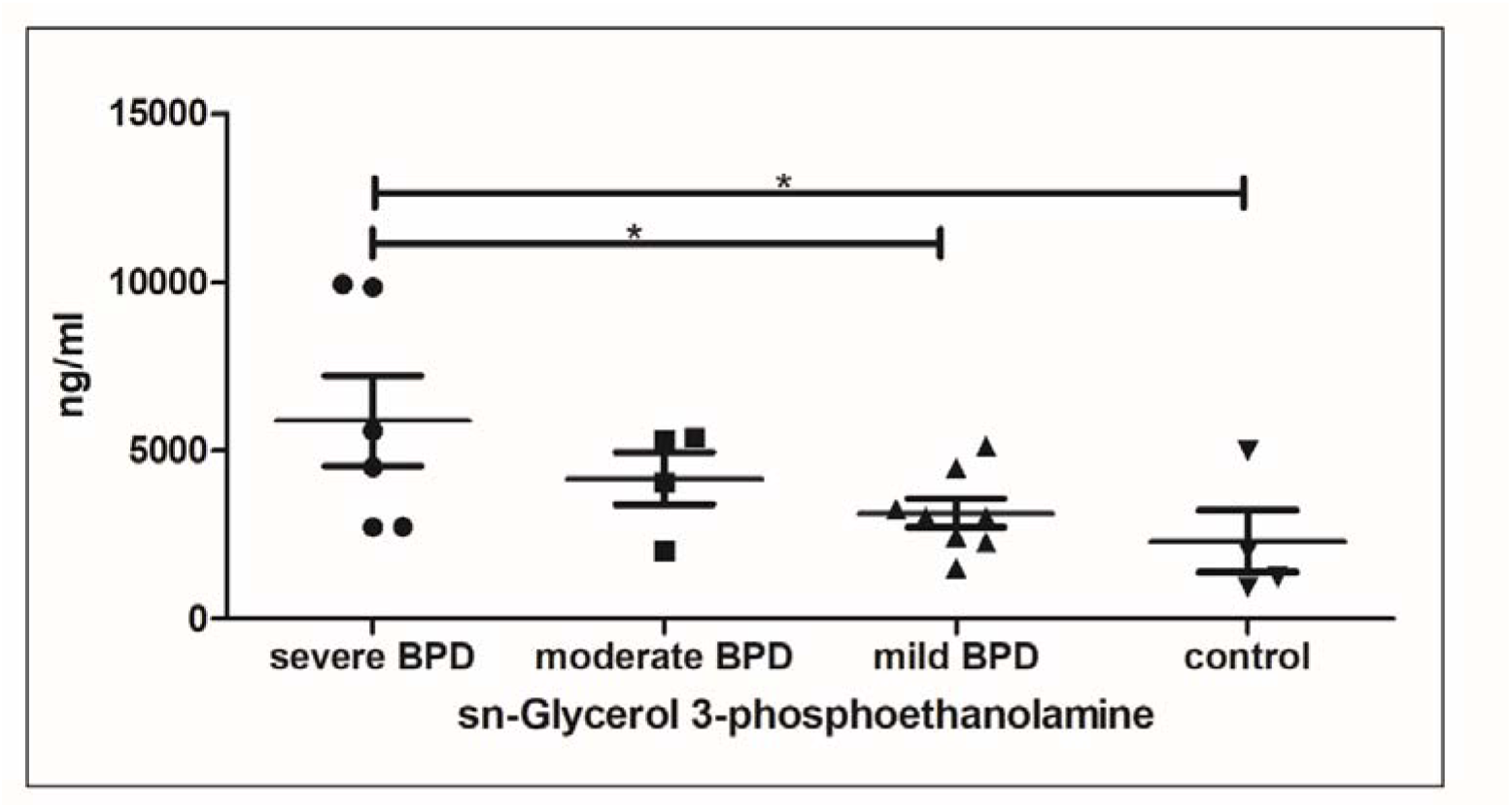
sn-Glycerol 3-phosphoethanolamine level at birth (Day 1) *:*P*< 0.05.

### 3.4 Correlation between the lower-airway microbiome and metabolites

Pearson’s correlation coefficient was used to explore the functional correlation between the changes in the lower-airway microbiome and differences in metabolites across the four groups at birth (Day 1) (**Figure 5**). There was a significant positive correlation between the abundance of *Stenotrophomonas* and sn-glycerol 3-phosphoethanolamine levels (r = 0.45) (P< 0.05).

**Figure 5:**
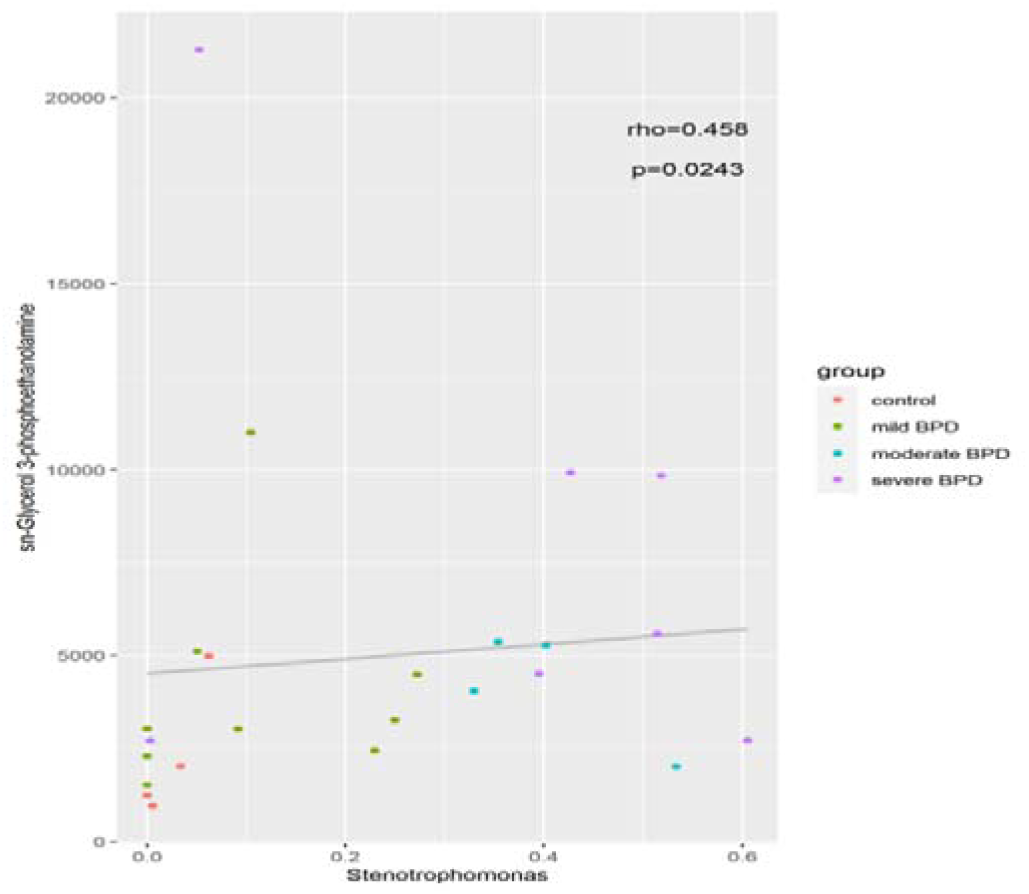
Scatter plot of the association between abundance of *Stenotrophomonas* and sn-glycerol 3-phosphoethanolamine level. r = 0.45, *P*< 0.05.

## Discussion

Studies investigating the correlation between BPD and the lower-airway microbiome in infants are scarce, and there remains an unmet clinical need to describe the lower-airway microbial communities and lower-airway metabolomic signatures in patients with BPD. Findings will allow the identification of microbial biomarkers for early detection of BPD and further understanding of the pathophysiology of BPD.

The present study used16S rRNA gene sequencing, metabolomic analyses, and the KEGG database in attempt to fill these evidence gaps. Results showed that multiple bacterial taxa can be identified in the respiratory secretions of intubated premature infants, even at birth and prior to surfactant administration. The diversity and composition of the lower-airway microbiome in infants with and without BPD varied at birth, but the differences became less pronounced on Day 7 of life. Consistent with a previous report, our study showed lower diversity in the lower-airway microbiome of infants that developed BPD [14]. Our results also revealed that alpha diversity of lower-airway communities was negatively correlated with BPD severity. At the phylum level, there were no significant differences in the composition of the lower-airway microbiome in infants with and without BPD or according to BDP severity. *Proteobacteria* was the most abundant microbe in all infants. These data align with one previous report, [14], but differ from another, which found that *Staphylococcus* and *Ureaplasma* were the most dominate lower-airway microbes in infants in their institution [10]. These disparate findings may be due to different environments, which likely influenced the composition of the lower-airway microbiome. At the genus level, *Stenotrophomonas* was significantly more abundant in infants with BPD compared to no BPD, and the abundance of *Stenotrophomonas* was positively correlated with BPD severity. These findings suggest that decreased diversity of the lower-airway microbiome and increased abundance of *Stenotrophomonas* in the lower-airway microbial community of intubated premature infants have potential as microbial biomarkers for early detection of BPD.

*Stenotrophomonas* is a nosocomial opportunistic pathogen of the *Xanthomonadaceae* family [17]. *Stenotrophomonas* isolated from the environment or in the clinical setting exhibits resistance to antibiotics and stress and forms biofilms on various surfaces, including the abiotic surfaces of catheters and prosthetic devices [18, 19]. *Stenotrophomonas* colonizes the lungs of patients with cystic fibrosis and those who are immunocompromised, and may represent a marker of chronic lung disease [18, 20–22]. *Stenotrophomonas* can influence the spatial organization and thus the function and composition of complex microbiomes [23, 24]. Data from the present study suggest a role for *Stenotrophomonas* in the pathogenesis of BPD in intubated premature infants.

As the composition of the lower-airway microbiome in preterm infants varied with the presence or absence of BPD and with BPD severity, we characterized the lower-airway metabolome in these infants. Findings showed significant differences between infants in 63 metabolites at birth (Day 1) and 29 metabolites on Day 7 of life, implying that metabolite variation paralleled that of the lower-airway microbiome. Among these metabolites, sn-glycerol 3-phosphoethanolamine was positively correlated with BPD severity, identifying it as a potential metabolic biomarker for early detection of BPD. The KEGG database showed that sn-glycerol 3-phosphoethanolamine has a role in glycerophospholipid metabolism. Glycerophospholipid has structural functions in bacteria, facilitates bacterial adaptation to environmental conditions, and is involved in bacteria–host interactions [28]. Glycerophospholipid is also associated with the pathophysiology of chronic obstructive pulmonary disease (COPD) [29]. These findings suggest that sn-glycerol 3-phosphoethanolamine may affect lower-airway microbiome composition and respiratory health in preterm infants. Our findings also identified a significant positive correlation between the abundance of *Stenotrophomonas* and sn-glycerol 3-phosphoethanolamine levels, an association that was confirmed with the KEGG database. This indicates that *Stenotrophomonas* may be abundant in the lower-airway microbiome of infants with BPD and responsible for the production of sn-glycerol 3-phosphoethanolamine, which may act as a pathogenic signal in these patients.

To the author’s knowledge, this prospective study is the first to identify a microbiome-metabolome signature in preterm infants with BPD. However, this study had several limitations. First, the sample size was small. Second, the data did not provide evidence that the lower-airway microbiome directly contributed to BPD. Although it will be challenging to determine a causal relationship between the lower-airway microbiome, metabolites and BPD development, further investigations are warranted.

In summary, there were significant differences in the diversity and composition of the lower-airway microbiome and metabolome in preterm infants with severe, moderate, or mild BPD or no BPD; the differences were more pronounced at birth (Day 1) than on Day 7 of life. Decreased diversity of the lower-airway microbiome, increased abundance of *Stenotrophomonas*, and increased level of sn-glycerol 3-phosphoethanolamine were positively associated with BPD severity, and have potential as predictive biomarkers for BPD. *Stenotrophomonas* may contribute to the development of BPD and influence the composition of the lower-airway microbiome through its metabolite, sn-glycerol 3-phosphoethanolamine. These findings provide novel insights into the lower-airway microbiome and its functions in BPD.

## Data Availability

All data referred to in the manuscript is available.

